# A survey of UK nurses about their care of people with malignant fungating wounds

**DOI:** 10.1101/2023.12.08.23299665

**Authors:** Susy Pramod, Jo Dumville, Gill Norman, Jacqui Stringer

## Abstract

**Background:** Malignant fungating wounds (MFW) are nonhealing wounds affecting people with advanced cancer. There is currently no research evidence on who delivers care for people’s MFW in the UK and what care these professionals deliver. This survey focussed on nurses who deliver care for people with MFW to find out more about their roles and the care they deliver, and to explore perceived barriers and facilitators to delivering care for these wounds.

**Method:** An online anonymous survey was conducted among nurses who provide wound care for people with MFW across the UK. Study data were collected using Qualtrics XM software and analysed with SPSS.

**Result:** We received 154 questionnaire responses. Respondents were tissue viability nurses, community nurses and other specialist nurses. The main reported MFW-related management aims were to manage wound odour, exudate, pain and bleeding, and prevention of infection. The top-ranked treatment aim was pain management followed by odour management. The most reported antimicrobial dressing was topical silver, and the non-antimicrobial dressing was superabsorbent. Access to MFW care training is reported as a barrier to providing care to people with MFW as is a lack of local and national guidelines. Availability of dressings, access to training, and good communication processes were reported as facilitators.

**Conclusion:** This is the first study that explored MFW wound care practices in the UK. A range of nurses are involved in care delivery with variations in the treatments used. Lack of access to MFW care training, resources, and standardised guidelines may impede care delivery.

## Introduction

MFW (MFW) are nonhealing wounds affecting people with advanced cancer. These wounds are caused by the aggressive proliferation of malignant cells and tumours that infiltrate the epidermis, blood vessels, and underlying structures (Grocott et al. 2013). These wounds predominantly develop during the last months of life and can indicate the impending end of life (Alexander 2010). MFWs can result from primary, secondary, or recurring malignant disease (Alexander 2009). A systematic review of studies from 1995-2020 by Tilley et al. (2021) found that MFWs can develop from any type of malignancy; the most prevalent are associated with breast cancer, followed by head and neck tumours. Malignancies of the groin, genitals, and back and other sites were also noted to lead to MFW. The review found that MFW can impact on the quality of life.

The epidemiology of fungating wounds in people with advanced cancer in the UK is limited. Data from a UK point prevalence study estimates that 1.47 people per 1000 have complex wounds at any one point (Gray et al. 2018), with the point prevalence estimate for MFWs being 0.02 per 1000 population (Hall et al. 2014). A survey of 269 nurses conducted in Switzerland in three different geographical regions over six months estimated that the prevalence of malignant wounds in patients with metastasised cancer was 6.6% (Probst et al. 2009). However, this was a survey of nurses and not a prospective prevalence study and so should be interpreted with caution. Prevalence data on people with MFWs from lower- and middle-income countries are not available in the published literature.

Malignant fungating wounds can be odorous, produce copious amounts of exudate (liquid from the wound), bleed easily, cause psychosocial distress and are a constant reminder to the patient and family of progressive cancer (Probst et al.2009). It is suggested that MFWs often become infected, although data on this are limited (Vardhan et al. 2019). A review exploring the MFW-related symptoms experienced by people (reporting data from 270 people) identified wound-related pain, malodour, exudate, bleeding, pruritus, and lymphedema (Tilley et al. 2020). The symptoms and symptom characteristics attributed explicitly to MFWs are unique to this population (Tilley et al. 2016).

Wound care for people with MFWs in several countries, including the UK, is normally the responsibility of nursing staff. There is limited data on care pathways for wound focused care for people with MFW, nor on the types of treatments that are widely being used. Topical treatments and dressings are likely to be the cornerstone of treatment regimes, although existing research evidence on the relative clinical or cost effectiveness is limited (Adderley and Holt 2014). The lack of research evidence in this field has been recognised to mean people with MFW are managed using practices based on anecdotal evidence, case studies and expert opinion (Gibson and Green 2013). This is still the case with more recent recommendations from European Oncology Nursing Society (EONS 2015).

Given the limited evidence base for the care of MFW, the care of people with these wounds has the potential to be heterogeneous. Yet, there is currently no reported data on who is delivering care for people’s MFW in the UK, what care they are undertaking and their insights into this. This means we have limited insights into service provision for people with MFW and staff views about this. In the UK, most people with MFW will likely be treated by community nurses; although care may also be received in hospices and via palliative care nurses, this has not been systematically explored. Tissue viability nurses offer specialist care in this area but may not see all people with MFW, depending on local care pathways and adherence to them. This survey focussed on nurses who deliver care for people with MFW to find out more about their roles, the care they deliver and to start to explore perceived barriers and facilitators to delivering this type of wound care.

The aim of the survey was:

- To gain insights into the nursing health professionals delivering care to people with MFW in the UK, where this care takes place and current referral practices.
- To understand health professionals’ wound-related treatment aims for people with MFW.
- To explore the wound care treatments used in the management of people with MFW and variations in their use.
- To explore health professionals’ confidence in delivering wound-related care to people with MFW and barriers and facilitators to delivering care.

## Methods

### Questionnaire design and piloting

An online anonymous questionnaire was developed to explore the activities and insights of wound care practitioners’ delivering care to people with MFW using convenience sampling from the UK. The questionnaire was constructed in QualtricsXM (Qualtrics, Provo, UT) following a process of question development and refinement. The questions were pretested for salience, flow, acceptability, administrative ease, and time taken to complete by piloting among expert wound care nurses (tissue viability nurses working in secondary care n=2 and in community care n=2, a district nurse (n=1) and a hospice nurse (n=1)). Amendments to the survey highlighted during the piloting phase were made prior to national distribution.

Questions formats included closed-form questions with single and multiple-choice options and free-text responses.

### Questionnaire Participants and Setting

The questionnaire link was sent to UK nurses who may deliver wound care. This included various nursing specialities: tissue viability nurses; district nurses; practice nurses; treatment room nurses, hospice-based nurses and oncology nurses. Recruitment was undertaken via professional organisations including regional and national nursing bodies and groups. The survey link was also circulated to relevant groups on Facebook and via Twitter. The survey was open for responses for a four-week period.

Participants were made aware that the purpose of this research is to gain an understanding into: who delivers care to people with MFW in the UK, where this takes place, current activities, staff confidence in providing care to people with MFW and any barriers and facilitators to care delivery they perceive. Individuals were eligible to participate in the survey if they:

- Were qualified nurses who have experience in wound care, including district nurses (i.e., those with a formal community nursing qualification and those with general qualifications working in the community), nurses working in oncology settings including hospices, treatment room nurses, practice nurses, and acute care nurses.
- Had a role in providing hands-on care for people with MFW within the participating clinical services.
- Cared for people with MFW in the last five years within the participating clinical services.

This study was reviewed by the University of Manchester Research Ethical Committee and was confirmed that ethical approval was not required due to the routine nature of the questions and their relation to clinical practice, the routes of staff recruitment and the anonymised data collection (letter available on request).

## Data analysis

Responses to the survey were recorded and summarised using QualtricsXM online software. Study data were analysed by using the SPSS Version 28.01.0 (142) (Statistical Package for Social Sciences). Descriptive statistics are used to present numerical data. Binary and categorical variables are summarised as percentages. Continuous data were summarised using mean and standard deviation and the median value with interquartile range. Where data are missing for binary and categorical fields, the denominator used in calculations is the total responses received for that question. Free text responses were summarised using a thematic approach. Information provided through the free-text comment section was read, collated and checked by the team members.

## Survey Result

### Respondents and their care for people with MFW

In total, we received 154 questionnaire responses that had one or more question response fields completed and thus were included in the analyses. We included responses from people who noted they did not **currently** deliver care to people with MFW, given that responses could relate to previous experiences within the last five years as per the criteria given for participation.

Three-quarters of respondents were tissue viability nurses (TVN), nearly 10% were community nurses and 13.7% were other specialist nurses which included palliative care nurses, oncology nurses, skin cancer specialists, wound care specialists, plastic surgery nurses, ward managers, and practice nurses (Table 1). Respondents delivered care to people with MFWs across a range of settings, most common of which were: community (55.2%), inpatient (49.4%); hospice (20.8%) and treatment rooms (19.5%) (Table 1). In total, 83 (53.9%) respondents reported delivering MFW care across more than one setting. Almost all respondents reported that they treated three or fewer people with a MFW at any one time and nearly half only treated one person with MFW at any one time (Table 1). There was variation in the frequency of reported patient contact for MFW-related care: 46.4% of respondents reported contact weekly or more frequently (Table 1).

**Table 1:** Summary of respondents’ role, setting(s) of care delivery, patient numbers and frequency of visit.

| **Clinical setting(s) treating people with malignant fungating wound\*** | **n (%) n=154** |
| --- | --- |
| Community | 85(55.2) |
| Hospital inpatients | 76(49.4) |
| Hospice | 32(20.8) |
| Treatment room | 30(19.5) |
| Hospital outpatients | 27(17.5) |
| Other care settings | 7(4.5) |
| *Missing data (not included on the denominator)* | 0 |
| **Role of health care professional caring for people with malignant fungating wound** | **n (%) n=153** |
| Tissue viability Nurse | 118(77.1) |
| Other specialist nurses | 21(13.7) |
| Community Nurses | 14(9.2) |
| *Missing data (not included on the denominator)* | 1 |
| **Number of people with malignant fungating wound treated at any one time by health care professionals** | **n (%) n=142** |
| 1 | 67(47.2) |
| 2 | 30(21.1) |
| 3 | 24(16.9) |
| 4 | 2(1.4) |
| 5 | 1(0.7%) |
| Others (varies from 1-4 or depends on the referral) | 8(5.6%) |
| *Missing data (not included on the denominator)* | 25 |
| Mean (Standard deviation) | 2.25(1.66) |
| Median (Inter Quartile Range) | 2(5) |
| **Frequency of reviewing a person with malignant fungating wound for care by health care professionals** | **n (%) n=140** |
| More than once a day | 5(3.6) |
| Daily | 14(10.0) |
| Three times a week | 4(2.9%) |
| Twice a week | 14(10.0) |
| Once a week | 28(20.0) |
| 1-2 weeks | 1(0.7%) |
| Once every two weeks | 28(20.0) |
| Monthly | 2(1.4%) |
| 6-8 weeks | 1(0.7%) |
| For initial assessment and plan only | 22(15.7) |
| Depending on needs | 21(15.0) |
| *Missing data (not included on the denominator)* | 14 |

\*responses do not sum to 100% as more than one setting could be selected

### Patient referral pathways

Respondents were asked who normally referred people with MFWs to them (Table 2). Over 60% of responding TVNs and community nurses reported receiving referrals from ward nurses. Over half of TVNs also reported receiving referrals from community nurses and a third reported getting referrals from palliative care and hospice nurses. Across all the nursing groups responding to the survey, referrals were received from a broad range of health and care professionals as well as patients themselves in a small number of cases.

**Table 2:** Referral source by respondent role for managing people with malignant fungating wound. *Referrals received from multiple sources.

| Role of respondent receiving the referral for managing people with malignant fungating wound |  |  |  |  |
| --- | --- | --- | --- | --- |
| Source of referral | Tissue viability nurse<br>n (%)<br>(n = 118) | Community nurse<br>n (%)<br>(n=13) | Another specialist<br>n (%)<br>(n=12) | Other<br>n (%)<br>(n=10) |
| Ward nurses | 79(66.9) | 8(61.5) | 6(50.0) | 5(50.0) |
| Community nurse | 69(58.5) | 1(7.7) | 3(25.0) | 5(50.0) |
| Palliative Care nurse | 48(40.7) | 5(38.5) | 5(41.7) | 7(70.0) |
| Patient themselves | 12(10.2) | 4(30.8) | 2(16.7) | 2(20.0) |
| Hospice nurse | 42(35.6) | 4(30.8) | 3(25.0) | 2(20.0) |
| Oncology nurse | 39(33.1) | 5(38.5) | 4(33.3) | 2(20.0) |
| Macmillan nurse | 23(19.5) | 3(23.0) | 3(25.0) | 3(30.0) |
| Other specialist nurses | 17(14.4) | (0) | 1(8.3) | 1(10.0) |
| Social worker | 3(2.5) | (0) | (0) | (0) |
| Informal carer | 2(1.7) | 4(30.8) | 1(8.3) | 1(10.0) |
| Missing data<br>(*not included on the denominator) | 1 | 1 | 1 | 1 |

*\*responses do not sum to 100% as referrals could be from multiple sources*

Respondents were then asked where they themselves refer patients with MFW on to for further specialist wound care. Respondents reported that they referred on average about one-third of the people with MFW in their care (35.5%).

Echoing data from Table 2, most onward referrals were to TVNs (Table 3). Nearly one-third (34.2%) of the TVNs did not refer their patients onwards, which is not unsurprising as they are wound specialists (Table 3). Where TVNs did refer patients onwards for wider support this included to pain teams, breast clinical nurse specialists, dermatologists and oncologists. When respondents were asked about the reason for referral to a specialist, the most frequent reason given was the wound becoming unmanageable (40.8%) (Table 4).

**Table 3:** Referral to by respondents for further specialist care. *Referrals sent to multiple destinations.

| Referral to for further specialist wound care (n=143) |  |  |  |  |  |  |  |  |
| --- | --- | --- | --- | --- | --- | --- | --- | --- |
| Respondent referring on | Comm<br>unity<br>nurses<br>n (%)<br>n=12 | Hospice<br>trained<br>nurses n<br>(%) n=2 | McMillan<br>nurses<br>n (%)<br>n=4 | Oncolo<br>gy<br>nurses<br>n(%)<br>n=10 | Palliativ<br>e care<br>nurses n<br>(%)<br>n=13 | Tissue<br>viability<br>nurses<br>n (%)<br>n=27 | Do not<br>refer<br>on<br>n (%)<br>n=46 | Others<br>n (%)<br>n=27 |
| **Tissue**<br>**viability**<br>**nurse (n =**<br>**111)** | 10(9.0) | 2(1.8) | 3(2.7) | 9(8.1) | 12<br>(10.8) | 11<br>(9.9) | 38<br>(34.2) | 26<br>(23.4) |
| **Community**<br>**nurse (n =**<br>**12)** | 0 | 0 | 1(8.3) | 0 | 0 | 11(91.7) | 0 | 0 |
| **Another**<br>**specialist**<br>**(n=11)** | 0 | 0 | 0 | 0 | 0 | 4(36.4) | 5(45.5) | 1(9.1) |
| **Other (n=9)** | 2(22) | 0 | 0 | 1(11.1) | 0 | 3(33.3) | 3(33.3) | 0 |
| **Missing**<br>**data (not**<br>**included on**<br>**the**<br>**denominator)** | 11 | 11 | 11 | 11 | 11 | 11 | 11 | 11 |

\*responses do not sum to 100% as referrals could be sent to multiple sources

**Table 4:** Reason for making a referral for specialist malignant fungating wound care.

| When do you refer a patient with malignant fungating wound for specialist wound care | n (%) n=142 |
| --- | --- |
| When the wound become unmanageable | 58(40.8%) |
| I do not refer these patients for further specialist wound care | 21(14.8%) |
| As soon as seen the patient | 12(8.5%) |
| When a patient asks | 10(7.0%) |
| When the wound is not improving | 9(6.3%) |
| When a patient is discharged | 9 (6.3%) |
| When symptom management is needed | 7(4.9%) |
| When the MDT approach is needed | 6(4.2%) |
| Tumour therapy is needed | 6(4.2%) |
| Depending on the patients' need | 2(1.4%) |
| After a certain period of time | 2(1.4%) |
| Missing data (*not included on the denominator) | 12 |

Participants were asked how long, in their experience, it took for a referred patient to be seen for specialist wound care and they reported that it took less than a week in most cases (54.4%) and most patients were seen within two weeks (17.2) (Table 5).

**Table 5:** Average time for reviewing a patient with malignant fungating wound following a referral by health care professionals.

| **The average time taken to review a patient with malignant fungating wound following a referral by health care professionals** | **n (%) n=93** |
| --- | --- |
| Less than a week | 51(54.8) |
| One week | 11(11.8) |
| Two weeks | 16(17.2) |
| Three weeks | 1(1.1) |
| Four weeks | 3(3.2) |
| Others | 11(11.8) |
| *Missing data (\*not included on the denominator)* | *61* |

### Management of MFW

Respondents reported that their main MFW-related management aims were to manage wound odour, exudate, and pain. Management of bleeding and prevention of infection were also common management aims. All the respondents identified all five areas as important in care (Table 6). Very few identified wound healing as a management aim, as would be expected since these wounds rarely heal or improve.

**Table 6:** Treatment aims when managing a person’s malignant fungating wound.

| **Treatment aims when managing a person's malignant fungating wounds\*** | **n (%) n=154** |
| --- | --- |
| Wound odour management | 143(92.6) |
| Exudate management | 142(92.2) |
| Pain management | 138(89.6) |
| Bleeding management | 129(83.8) |
| Prevention of infection | 122(79.2) |
| Healing | 5(3.2) |
| *Missing data* | *0* |

*\*responses do not sum to 100% due to the nature of the treatment aims*

Respondents were asked to rank these management aims (Table 7). Twice as many respondents ranked wound pain as their top management aim compared with any other options and three-quarters placed it in their top three priorities.

**Table 7:** Top rankings of malignant fungating wound management aims (n = 125)

| **Malignant fungating wound management aims** | **Ranked 1<sup>st</sup>**<br>**n (%)** | **Ranked 2<sup>nd</sup>**<br>**n (%)** | **Ranked 3<sup>rd</sup>**<br>**n (%)** | **Ranked 4<sup>th</sup>**<br>**n (%)** | **Ranked 5<sup>th</sup>**<br>**n (%)** |
| --- | --- | --- | --- | --- | --- |
| Pain management | 58(46.4) | 25(20.0) | 19(15.2) | 15(12.0) | 8(6.4) |
| Wound odour management | 25(20.0) | 22(17.6) | 29(23.2) | 25(20.0) | 24(19.2) |
| Bleeding management | 20(16.0) | 24(19.2) | 33(26.4) | 23(18.4) | 25(20.0) |
| Exudates management | 10 (8.0) | 39(31.2) | 37(29.6) | 31(24.8) | 8(6.4) |
| Prevention of infection | 7 (5.6) | 18(14.4) | 10(8.0) | 30(24.0) | 60(48.0) |
| Healing | 6(4.8) | 1(0.8) | 1(0.8) | 2(1.6) | 115(92.0) |
| *Missing data (\*not included on the denominator)* | 29 | 29 | 29 | 29 | 29 |

### Treatments for MFW

Participants were asked about the different types of treatments commonly used for MFW care under the categories of dressings (antimicrobial and non-antimicrobial); and medications (systemic antibiotics, types of steroids and other products) (Figure 1).

**Figure 1:**
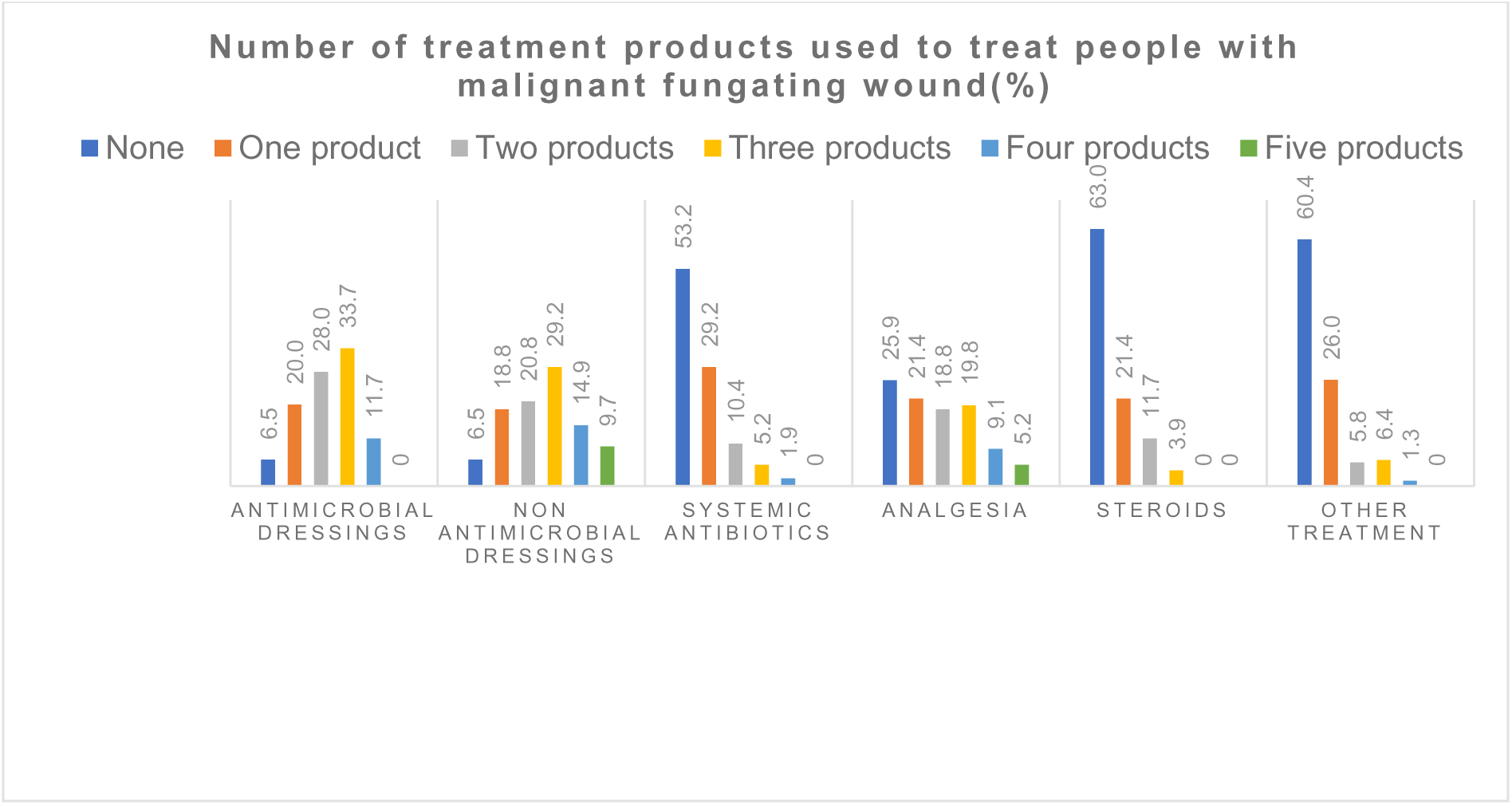
Number of products used to treat people with malignant fungating wound.

### Dressings

In total, 93.5% of respondents reported using antimicrobial dressings and topical agents for the treatment of people with MFW (Figure 1). In total 64% of respondents reported using topical silver within this category, and over half of the respondents reporting using topical metronidazole (55.2%) (Figure 2). Respondents also frequently reported the use of non-antimicrobial dressings (93.4% of respondents reported using at least one non-antimicrobial dressing) (Figure 1). Respondents most frequently reported the use of superabsorbent dressings (79.2%) (Figure 2).

**Figure 2:**
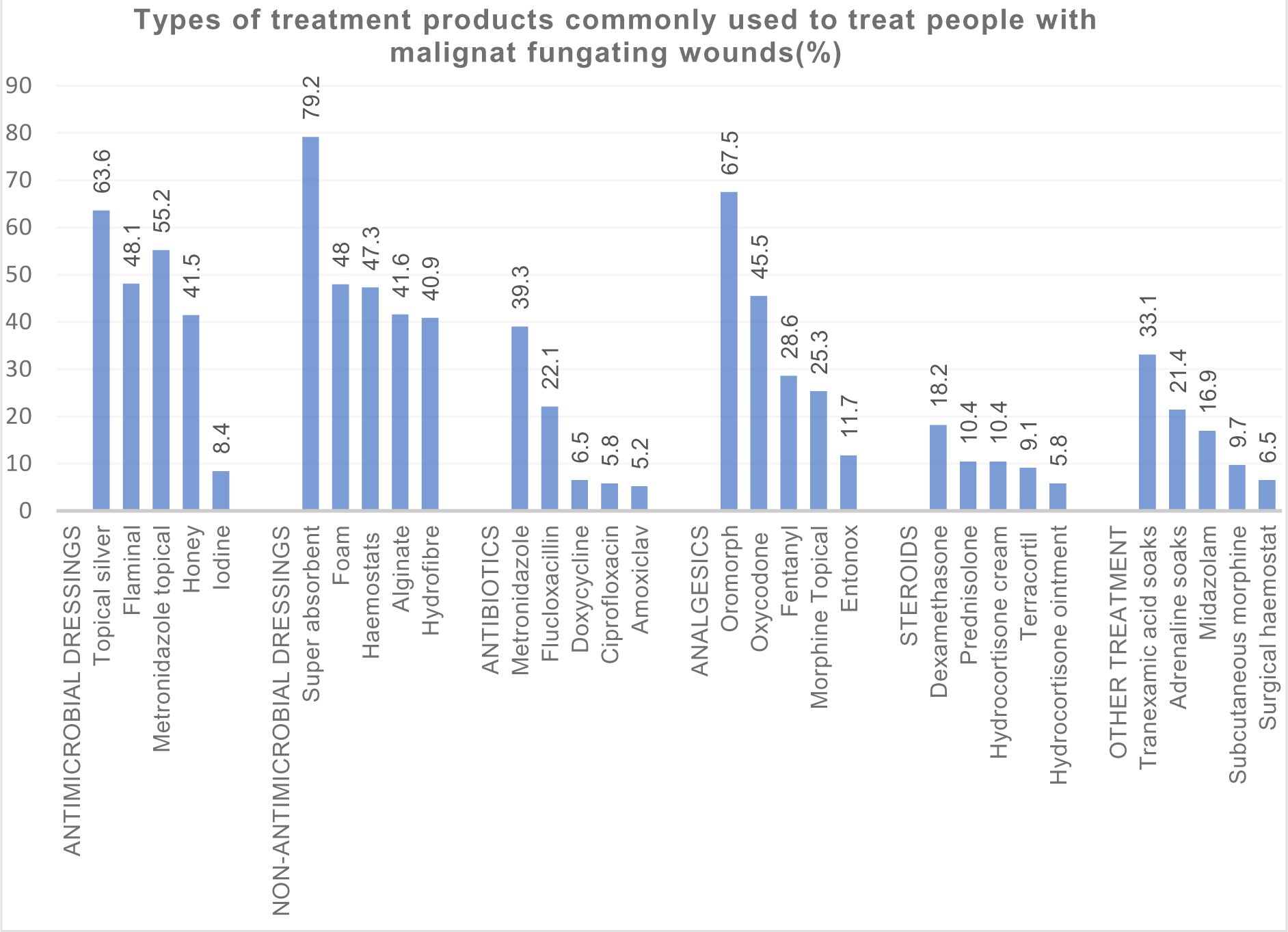
Types of treatment products commonly used to treat people with malignant fungating wounds.

### Medications

Use of one or more types of systemic antibiotics for managing MFW was reported by less than half of respondents (46.7%) (Figure 1) with Metronidazole the most frequently reported product (39.3%) (Figure 2). In total, 25.9% of the respondents reported they do not commonly use any analgesia to manage MFW (Figure 1). The most commonly used analgesia reported was Oromorph^(™)^ (67.5% of respondents reported use) (Figure 2). Steroid use was less frequently reported with 67.5% of respondents reporting they do not use any type of steroids (Figure 1). The most used other treatments for MFW were reported as Tranexamic acid soaks (33.1%), Adrenaline soaks (21.4%) and Midazolam (16.9%) (Figure 2).

### Delivery of MFW care

We also asked about the participants’ confidence in providing care to people with MFW. Three-quarters of the TVNs were very confident or confident in providing care to people with MFW (Table 8). The people with no confidence or limited confidence were in a minority and from different banding and job roles.

**Table 8:** Level of confidence in caring for people with malignant fungating wound.

| The confidence level of health care professional in caring for people with malignant fungating wound | Tissue viability nurses n (%) n=108 | Community Nurses n (%) n=12 | Another specialist nurse n (%) n=12 | Others n (%) n=9 | Overall response n (%) n=141 |
| --- | --- | --- | --- | --- | --- |
| Very confident | 24(22.2) | 0 | 2(16.7) | 1(11.1) | 27(19.1) |
| Confident | 49(45.4) | 6(50.0) | 5(41.7) | 8(88.9) | 68(48.2) |
| Somewhat confident | 25(23.1) | 5(41.7) | 4(33.3) | 0 | 34(24.1) |
| Limited confidence and no confidence | 10 (9.3) | 1(8.3) | 1(8.3) | 0 | 12(8.5) |
| Missing data (*not considered on the denominator) |  |  |  |  | 13 |

### Barriers and facilitators of care provision for MFW

Over half of the respondents (51.9%) reported that access to training on delivering MFW care is a barrier in delivering wound care to this patient group and 24.7% reported access to training facilitated in providing care (Figure 3). Communication between care settings was reported as a barrier by 31.2% of respondents and 34.4% respondents reported communication between the care setting facilitated in providing care. Availability of dressings was a barrier (26.0%) in providing care and 46.1% reported availability of dressings facilitated care for people with malignant fungating wound.

**Table 9:** Existing barriers and facilitators in providing malignant fungating wound care (n = 154)

**Figure 3:**
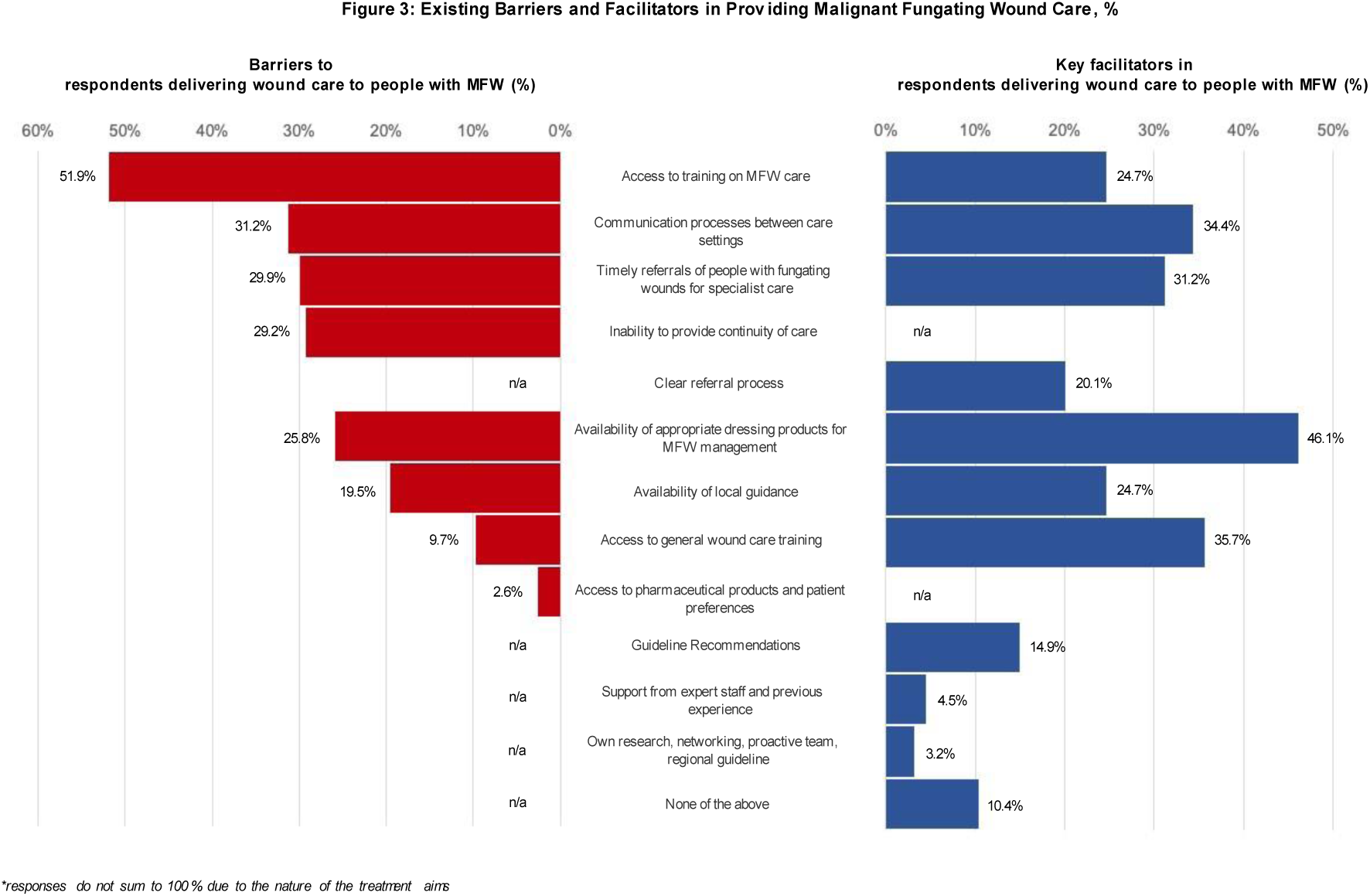
Existing Barriers and Facilitators in Providing Malignant Fungating Wound Care %.

In the questionnaire we asked whether respondents had access to local guidance on the wound care of MFW, 54.6%respondents said yes (36.9% no, 8.5% didn’t know and 13 missing data from n = 154). Almost a fifth of respondents (19.5%) cited a lack of local guidance as a barrier to care delivery. When asked about any further comments on MFW care delivery,25% (n=44) of the respondents reported a lack of national guidelines as a barrier

## Discussion

### Staff delivering wound care to people with MFWs

This survey is the first in the UK to explore current UK practices in caring for people with MFW. Whilst the survey was distributed widely, the majority of responses were from tissue viability nurses, who in the UK are specialist wound care nurses and could be expected to be responsible for complex wounds such as MFWs (Ousey et al. 2014).

Survey findings confirm that tissue viability nurses (TVN) are often a referral point from the community and other nursing staff, and these specialist nurses largely reported confidence in providing care to people with MFWs. However, there was variation in confidence levels within the tissue viability group. This may reflect the relatively small number of people with MFW relative to other wound types and lack of correct differential diagnosis, experience and confidence. Respondents reported a lack of training as the main barrier to delivering care to people with MFW, others supporting this by recognising training as a facilitator of care provision. Inadequacies in wound care training for nurses at all levels has been reported previously (Welsh 2018). Other studies have highlighted that barriers to training in wound care can be related to nurses’ ability to access training due to busy workloads and lack of capacity as well as staff turnover rates (Perry et al. 2022). Furthermore, practical training and education for those who treat non-healing wounds have not changed despite advances in wound care and dressing technology (Atkin 2020).

### Management of malignant fungating wounds

Management of MFWs poses a significant challenge to health care professionals and in most cases, the aim is to manage the symptoms rather than healing (Tandler and Stephen-Haynes 2017).

Pain in advanced cancer is one of the most common and distressing symptoms reported (Tilley et al. 2016) and management of wound-related pain was a high priority MFW symptom for management reported by respondents. As people with MFW will experience wound and cancer related symptoms simultaneously they may experience different types of pain including deep pain, neuropathic pain, and superficial pain related to procedures(Starace et al. 2022). It is reported that the mechanism involved in MFW-related pain is complex and multi-factorial because of the omnipresence of infection due to high local microbial load (Vardhan et al. 2019), irritation of exposed nerve endings (trauma at dressing changes), tumour growth (pressure compressing on body structures) and oedema (impaired lymphatic and capillary drainage) (da Costa Ferreira et al. 2023).

Whilst pain management was most commonly reported as the highest priority aim of wound care, a quarter of respondents reported that they do not commonly use any analgesia in this patient group. There are a number of possible reasons for this: a previous study has noted that wound-related pain generally can be under-assessed and may therefore be sub-optimally managed (Frescos 2018). It may be a knowledge or training issue around MFW-related pain assessment or management (Nuseir et al. 2016). Or there may be other reasons some nurses are not reporting use of MFW-related analgesia such as patients being on other cancer-related analgesia, and therefore further pain relief not being required. However, it is important to acknowledge that pain associated with chronic wounds can affect quality of life, and has a major impact on physical, emotional, and cognitive function (Frescos 2018).

In the survey, odour management was ranked as the second most important treatment aim in managing people with MFW. Wound-related odour has previously been reported by health professionals and patients as a main symptom associated with MFWs (Gethin et al. 2014) and as being both a principal source of patient distress and a challenge in wound management (Gibson and Green 2013). Several studies reported evidence on and use of charcoal dressings, silver dressings and topical metronidazole in the management of odour in people with MFW (da Costa Santos et al. 2010; Gethin et al.2014; de Castro and Santos 2015; Tsichlakidou et al. 2019; Gethin et al. 2023; O’eill et al. 2022). However, there is limited evidence to support the effectiveness of these approaches for the management of odour (Finlayson et al. 2017).

### Treatment use and available evidence

In terms of treatments used by respondents to deliver wound care to people with MFW, of note was the commonly reported use of antimicrobial dressings. The product mostly used within this class was silver dressings with topical metronidazole also commonly used. Whilst somewhat dated, systematic review evidence has highlighted a lack of robust comparative effectiveness evidence to guide decision making about the use of topical treatments and dressings (Adderley and Holt 2014) and topical antimicrobials (Finlayson et al. 2017) in MFW management. More widely, there is currently no clear evidence that silver-containing dressings confer clinical or cost effectiveness for wounds relative to other treatment options on outcomes such as healing, treatment and prevention of infection and odour management. These dressing are not recommended in NICE Guidelines (2016) for treatment of complex wounds although they have been widely used (Hussey et al. 2019). We do note however, that silver dressings are mentioned for use by a NICE Clinical Knowledge Summary (NICE CSK 2023) around infected wound that considered care in people with MFW.

The common use of silver dressings reported in this survey for MFW care reflects a perception that wound odour and/or pain is related to increased bacterial burden or infection (de Castro and Santos 2015) and that silver-containing dressings may reduce bacterial load and thus symptoms experienced by patients (Kalemikerakis et al. 2012). However, whilst we collected treatment data from respondents in the survey, we did not ask staff what motivated their treatment choice, so this requires further exploration. Interestingly, respondents noted the availability of appropriate dressings and products for MFW as a facilitator in providing MFW care. What dressings and other treatments are considered as appropriate for MFW care and how this relates to evidence, clinical experience and available responses is likely a valuable future area of study.

As well as limited evidence to inform decision making for treatment selection for people with MFW, we are not aware of any current national guidelines focused on MFW care. There are recommendations from European Oncology Nursing Society (EONS 2015) on Care of Patients with Malignant Fungating Wounds including how to assess and manage the symptoms associated with MFWs. In addition, evidence summaries such as NICE CKS on palliative care and malignant skin cancers deal with “how to manage a malignant skin ulcer” in terms of dressings choices for managing pain, odour, exudate, bleeding and infection, recommending an expert professional involvement in wound care to improve the quality of life of the people with MFW (NICE CKS 2023).

The survey found that some HCPs utilise local guidance or pathways to manage the symptoms of MFW. It is reported that customising guidelines to local conditions could weaken the integrity of the evidence base (Harrison et al. 2010). Even though local guidelines or pathways can improve decision making when uncertain, they can be highly influenced by the clinical experience and opinion of the developer which can misinterpret the evidence or may not be suitable for the individual patient (Woolf et al. 1999).

## Limitations

Although the survey was sent across the UK aimed at clinical staff who deliver MFW care, with efforts to reach hospice nurses, community nurses and practice nurses, most respondents were TVNs. The reason for the over-representation of TVNs in the survey is not clear. It could be due to the reach of the survey, or because TVNs make up the highest proportion of staff who deliver MFW care. We also note that the results of this survey will be most relevant to the UK and its health care system. This survey has not explored the level of professional qualifications of the participants in wound management, which can have an impact on the delivery of care to people with MFW. This survey only explored physical symptom management and did not explore the psychological aspect of care provided to people with MFW. Moreover, we have not examined the psychological issues of staff delivering MFW care. This has been explored by qualitative studies and indicates that the areas identified as priorities are experienced as most challenging since this affects the patient’s quality of life (Ousey et al. 2016). The survey only explored the management aim for treating MFW from professional’s viewpoint and did not explore the service user point of view. The experiences of patients with MFW and their carers have been explored qualitatively (Gibson and Green 2013) and should be considered alongside the findings of this survey.

## Strengths

To the best of our knowledge, this is one of the first studies to investigate the care of people with MFW by nurses in the UK. The survey only focussed on staff who currently care for or have cared for people with MFWs in the last five years, to collect the latest practices. This survey was anonymous, encouraging honest self-evaluation by respondents and the results were analysed using SPSS software which avoided manual extraction, errors and bias. Most questions had an “other” option for responses in case desired responses were not available from the drop-down menu.

## Conclusions and recommendations for future research

Based on the results of this survey, we gained insight into the experiences of nurses who deliver wound care to people with MFW. TVNs may be responsible for much of this care, although nurses from across services are likely to benefit from knowledge about treating people with MFWs.

There is variation in treatment use in MFW care, with a need for further evidence to support decision making. The development of guidelines in the interim may support more consistent decision making at a local level and allow recognition of uncertainties that may not currently be well recognised. This may also support access to treatments that, in the current absence of comparative evidence, represent the current consensus on the optimal use of available resources and clinical and patient acceptability.

Whilst we are not aware of recent substantive comparative studies of treatments for MFW, updating of existing systematic reviews will be important to inform decision making. Further exploration with staff and patients about what is driving use of treatments with weak evidence bases such as antimicrobial dressings for MFW care will add further insights. Pain management in people with MFW needs further research to fully understand the reasons for not utilising pain medication in spite of pain being a priority of care. It may be worthwhile to investigate whether staff experience and levels of confidence in providing care have any influence.

Currently, no information is available about who delivers training to staff to manage MFW or if training is appropriate and meets user needs. For clinical and financial impact on treatment decisions, clinical and cost-effectiveness research may be necessary. As this survey did not explore the experience from the patient perspectives, future research should focus on exploring the patient perspectives and impact on quality of life to understand what is important for them in their MFW management.

## Author contribution

Susy Pramod (SP), Jo Dumville (JD), Gill Norman (GN) and Jacqui Stringer (JS) conceived the idea for the overall project and contributed to the detailed development of the study design. SP collected the data and conducted the data analysis. SP, JD, GN, and JS contributed to the interpretation of study findings. SP wrote the first draft of the manuscript; JD, GN and JS revised and edited the manuscript for important intellectual content and all authors approved the final manuscript for submission.

## Data availability statement

Data are available on reasonable request. Requests for access to data should be addressed to the corresponding author.

## Data Availability

All data produced in the present study are available upon reasonable request to the corresponding author.

## References

Adderley, U.J. and Holt, I.G.S. 2014. Topical agents and dressings for fungating wounds. Cochrane Database of Systematic Reviews 2014(5). doi: 10.1002/14651858.CD003948.pub3.

Alexander, S. 2009. Malignant fungating wounds: epidemiology, aetiology, presentation and assessment. Journal of wound care 18(7), pp. 273–276.

Alexander, .J. 2010. An intense and unforgettable experience : the lived experience of malignant wounds from the perspectives of patients, caregivers and nurses. International Wound Journal 7(6) pp.456–65.

Atkin, L. 2020. Education in wound care: the need to improve access. Journal of General Practice Nursing 6(1), pp. 37–41.

De Castro, D.L.V. and Santos, V.L.C.G. 2015. Odor management in fungating wounds with metronidazole: A systematic review. Journal of Hospice and Palliative Nursing 17(1), pp. 73–79. doi: 10.1097/NJH.0000000000000125.

da Costa Ferreira, S.A., Serna González, C.V., Thum, M., da Costa Faresin, A.A., Woo, K. and de Gouveia Santos, V.L.C. 2023. Topical therapy for pain management in malignant fungating wounds: A scoping review. Journal of Clinical Nursing 32(13– 14), pp. 3015–3029. doi: 10.1111/jocn.16508.

da Costa Santos, C.M., de Mattos Pimenta, C.A. and Nobre, M.R.C. 2010. A Systematic Review of Topical Treatments to Control the Odor of Malignant Fungating Wounds. Journal of Pain and Symptom Management 39(6), pp. 1065– 1076. doi: 10.1016/j.jpainsymman.2009.11.319.

EONS 2015. Recommendations for the Care of Patients with Malignant Fungating Wounds. European Oncology Nursing Society, pp1–30.

Finlayson, K., Teleni, L. and McCarthy, A.L. 2017. Topical opioids and antimicrobials for the management of pain, infection, and infection-related odors in malignant wounds: A systematic review. Oncology Nursing Forum 44(5), pp. 626–632. doi: 10.1188/17.ONF.626-632.

Frescos, N. 2018. Assessment of pain in chronic wounds: A survey of Australian health care practitioners. International Wound Journal 15(6), pp. 943–949. doi: 10.1111/iwj.12951.

Gethin, G. et al. 2023. Systematic review of topical interventions for the management of odour in patients with chronic or malignant fungating wounds. Journal of Tissue Viability 32(1), pp. 151–157. Available at: 10.1016/j.jtv.2022.10.007.

Gethin, G., Grocott, P., Probst, S. and Clarke, E. 2014. Current practice in the management of wound odour: An international survey. International Journal of Nursing Studies 51(6), pp. 865–874. Available at: 10.1016/j.ijnurstu.2013.10.013.

Gibson, S. and Green, J. 2013. Review of patients’ experiences with fungating wounds and associated quality of life. Journal of Wound Care 22(5), pp. 265–275. doi: 10.12968/jowc.2013.22.5.265.

Gray, T.A., Rhodes, S., Atkinson, R.A., Rothwell, K., Wilson, P., Dumville, J.C. and Cullum, N.A. 2018. Opportunities for better value wound care: A multiservice, cross-sectional survey of complex wounds and their care in a UK community population. BMJ Open 8(3), pp. 1–9. doi: 10.1136/bmjopen-2017-019440.

Grocott, P., Gethin, G. and Probst, S. 2013. Malignant wound management in advanced illness : new insights., pp. 101–105. doi: 10.1097/SPC.0b013e32835c0482.

Hall, J., Buckley, H.L., Lamb, K.A., Stubbs, N., Saramago, P., Dumville, J.C. and Cullum, N.A. 2014. Point prevalence of complex wounds in a defined United Kingdom population. doi: 10.1111/wrr.12230.

Harrison, M.B., Légaré, F., Graham, I.D. and Fervers, B. 2010. Adapting clinical practice guidelines to local context and assessing barriers to their use. *CMAJ*. Canadian Medical Association Journal 182(2). doi: 10.1503/cmaj.081232.

Hussey, L., Stocks, S.J., Wilson, P., Dumville, J.C. and Cullum, N. 2019. Use of antimicrobial dressings in England and the association with published clinical guidance: Interrupted time series analysis. BMJ Open 9(9), pp. 1–9. doi: 10.1136/bmjopen-2018-028727.

Kalemikerakis, J., Vardaki, Z., Fouka, G., Vlachou, E., Gkovina, U., Kosma, E. and Dionyssopoulos, A. 2012. Comparison of foam dressings with silver versus foam dressings without silver in the care of malodorous malignant fungating wounds. Journal of B.U.ON. 17(3), pp. 560–564.

National Institute for Care and Excellence (NICE) 2016. Chronic wounds: advanced wound dressings and antimicrobial dressings. https://www.nice.org.uk/advice/esmpb2/resources/chronic-wounds-advanced-wound-dressings-and-antimicrobial-dressingspdf-1502609570376901 (Accessed September 2023).

National institute For Care and Excellence Clinical Knowledge Summary (NICE-CKS). 2023. Palliative care - malignant skin ulcer https://cks.nice.org.uk/palliative-care-malignant-skin-ulcer (Accessed October 2023)

Nuseir, K., Kassab, M. and Almomani, B. 2016. Healthcare providers’ knowledge and current practice of pain assessment and management: How much progress have we made? Pain Research and Management 2016, pp. 18–23. doi: 10.1155/2016/8432973.

O’Neill, L. et al. 2022. Malignant Fungating Wounds of the Head and Neck: Management and Antibiotic Stewardship. OTO Open 6(1). doi: 10.1177/2473974X211073306.

Ousey, K., Atkin, L., Milne, J. and Henderson, V. 2014. The changing role of the tissue viability nurse: An exploration of this multifaceted post. Wounds UK 10(4), pp. 54–61.

Ousey, K., Roberts, D. and Learning, C. 2016. Exploring nurses’ and patients’ feelings of disgust associated with malodorous wounds: a rapid review. 25(8), pp. 25–28.

Perry, C. et al. 2022. What promotes or prevents greater use of appropriate compression in people with venous leg ulcers? A qualitative interview study with nurses in the north of England using the Theoretical Domains Framework. BMJ Open 12(8). doi: 10.1136/bmjopen-2022-061834.

Probst, S., Arber, A. and Faithfull, S. 2009. European Journal of Oncology Nursing Malignant fungating wounds : A survey of nurses’ clinical practice in Switzerland. European Journal of Oncology Nursing 13(4), pp. 295–298. Available at: 10.1016/j.ejon.2009.03.008.

Starace, M. et al. 2022. Management of malignant cutaneous wounds in oncologic patients. Supportive Care in Cancer 30(9), pp. 7615–7623. Available at: 10.1007/s00520-022-07194-0.

Tandler, S. and Stephen-Haynes, J. 2017. Fungating wounds: Management and treatment options. British Journal of Nursing 26(12), pp. S6–S14. doi: 10.12968/bjon.2017.26.12.S6.

Tilley, C.P., Cleeve, J. Van, Crocilla, B.L. and Confort, C.P. 2020. and Functional Performance among Patients with Advanced Cancer : Journal of palliative medicine 23(6). doi: 10.1089/jpm.2019.0617.

Tilley, C.P., Fu, M.R., Qiu, J.M., Comfort, C., Crocilla, B.L., Li, Z. and Axelrod, D. 2021. The Microbiome and Metabolome of Malignant Fungating Wounds: A Systematic Review of the Literature from 1995 to 2020. *Journal of Wound*, Ostomy and Continence Nursing 48(2), pp. 124–135. doi: 10.1097/WON.0000000000000749.

Tilley, C.P., Lipson, J. and Ramos, M. 2016. Palliative Wound Care for Malignant Fungating Wounds Holistic Considerations at End-of-Life. Nurs Clin N am 51, pp. 513–531.

Tsichlakidou, A., Govina, O., Vasilopoulos, G., Kavga, A. and Kalemikerakis, I. 2019. Intervention for symptom management in patients with malignant fungating wounds - a systematic review. Journal of BUON 24(3), pp. 1301–1308.

Vardhan, M. et al. 2019. The Microbiome, Malignant Fungating Wounds, and Palliative Care. Frontiers in Cellular and Infection Microbiology 9(November), pp. 1–7. doi: 10.3389/fcimb.2019.00373.

Welsh, L. 2018. Wound care evidence, knowledge and education amongst nurses: a semi-systematic literature review. International Wound Journal 15(1), pp. 53–61. doi: 10.1111/iwj.12822.

